# Clinical determinants and outcomes associated with the development of acute kidney injury in critically ill patients: a Brazilian retrospective cohort study

**DOI:** 10.1101/2025.01.29.25321328

**Authors:** Rodrigo C. Menezes, Rodrigo S.F. Meira-Teles, Lucas G. Moura, Isabella B. B. Ferreira, Nivaldo M. Filgueiras Filho, Bruno B. Andrade

**Affiliations:** Laboratório de Pesquisa Clínica e Translacional, Instituto Gonçalo Moniz (IGM), Fundação Oswaldo Cruz (FIOCRUZ), Salvador, Brazil; Multinational Organization Network Sponsoring Translational and Epidemiological Research (MONSTER) Initiative, Salvador, Brazil; Instituto de Pesquisa Clínica e Translacional, Faculdade ZARNS, Salvador Brazil; Curso de Medicina, Escola Bahiana de Medicina e Saúde Pública, Salvador, Brazil; Intensive Care, Hospital da Cidade, Salvador, Brazil; Medicina, Universidade do Estado da Bahia (UNEB), Salvador, Brazil; Programa de Pós-Graduação em Medicina e Saúde Humana, Escola Bahiana de Medicina e Saúde Pública (EBMSP), Salvador, Brazil

**Keywords:** Acute kidney injury, Risk factors, Outcomes, Bayesian, Intensive care unit

## Abstract

**Background:** Acute kidney injury (AKI) is a common complication in intensive care unit (ICU) patients and associates with significant morbidity and mortality. Understanding region-specific risk factors is crucial for early identification and management, especially in resource-limited settings. This study aimed to identify characteristics, risk factors, and outcomes related to AKI in critically ill patients from a center in Northeast Brazil.

**Methods:** Retrospective cohort study that used secondary data of adult ICU patients admitted to a medium-sized hospital in Brazil between August 2015 and April 2024. Patients with pre-existing chronic kidney disease or AKI at admission were excluded. Demographic and clinical variables, documented comorbidities, clinical and laboratory data in the first 6 hours of admission, and adverse events during hospitalization were collected. To evaluate risk factors, the variables were included in a multivariable Cox regression model, while to assess complications, a univariable model was employed. Bayesian network modeling was applied to infer causal relations between the presence of AKI and the statistically significant risk factors. Kaplan-Meier curves were used to evaluate survival and length of stay.

**Results:** Of 5,416 patients analyzed, 369 (6.81%) developed AKI during their ICU stay. Independent risk factors for AKI included advanced age (adjusted Hazard Ratio [aHR] 1.01 per increase in 1 year of age; 95% CI: 1.00–1.02; p=0.008), chronic liver failure (aHR 10.02; 95% CI: 4.91–20.41; p<0.001), arterial hypertension (aHR 1.29; 95% CI: 1.00–1.66; p=0.048), higher heart rate at admission (aHR 1.01 per bpm; 95% CI: 1.00–1.01; p=0.004), and reduced level of consciousness (aHR 1.67 per point decrease in Glasgow Coma Scale; 95% CI: 1.34–2.07; p<0.001). Underweight patients had a lower risk of AKI compared to those with normal weight (aHR 0.59; 95% CI: 0.38–0.91; p=0.017). AKI was associated with higher mortality (37.13% vs. 10.54%; p<0.001) and longer hospital stay (median 16 vs. 10 days; p<0.001).

**Conclusions:** Age, chronic liver failure, arterial hypertension, higher heart rate, and reduced level of consciousness at admission are independent risk factors for AKI development. AKI was associated with poorer outcomes, highlighting the need for early identification and targeted interventions in this population.

## BACKGROUND

Acute kidney injury (AKI) is a common and severe complication among critically ill patients admitted to intensive care units (ICUs), with reported incidences reaching up to 50% of admissions [1–3]. Characterized by a sudden decline in kidney function, AKI disrupts fluid, electrolyte, and acid-base balance, and is associated with increased morbidity and mortality, prolonged hospital stays, and elevated healthcare costs [4,5]. Early identification of patients at risk for AKI is essential to facilitate timely diagnosis, avoid nephrotoxic exposures, and implement appropriate hemodynamic management [6,7].

Previous studies have identified important risk factors for AKI development, particularly advanced age, obesity, arterial hypertension, diabetes, heart disease, chronic liver failure and hypotension during ICU stay [8–14]. However, these studies often enrolled patients with pre-existing chronic kidney disease (CKD) or acute kidney injury at the time of admission, confounding the identification of risk factors specific to ICU-acquired AKI. Furthermore, most studies have evaluated risk factors in developed countries, and although the development of AKI has a uniform global incidence, regional outcomes may vary due to intrinsic clinical, sociodemographic, and epidemiologic factors [15–17].

In Brazil, changes in the epidemiological profile, including an aging population and a rise in chronic diseases, such as diabetes and hypertension, have likely contributed to an increase in AKI incidence [18]. Additionally, the country’s healthcare system is marked by regional disparities, with significant variations in ICU resources and management practices, potentially influencing AKI incidence and outcomes. Understanding AKI in this context can inform preventive strategies tailored to resource-constrained environments.

Therefore, this study aims to assess the clinical characteristics and risk factors associated with the development of AKI and the main outcomes in the ICU setting, seeking to improve patient prognosis through the early identification and the implementation of targeted management strategies.

## MATERIALS AND METHODS

### Study design and participants

Retrospective observational cohort study that included all adult patients (≥ 18 years old) admitted to the ICU of the Hospital Cidade in Salvador, Brazil, between August 2015 and April 2024. Patients with pre-existing CKD or with AKI at admission and those with incomplete baseline or outcome data were excluded.

### Data collection

Clinical data were obtained using the Epimed Monitor System, an electronic platform specifically designed for real-time patient monitoring in ICUs, at 06/05/2024. Daily, ICU nurses entered patient data into the system as part of routine care, following standardized local protocols to ensure consistency and accuracy.

Demographic and clinical variables collected at ICU admission included age, sex, body mass index (BMI), type of admission (medical, elective surgery, or emergency surgery), Simplified Acute Physiology Score 3 (SAPS 3), readmission status, hospital discharge outcome, and lengths of stay in the ICU and hospital. Documented comorbidities included cardiovascular, respiratory, metabolic, and other chronic conditions (e.g., heart failure, chronic obstructive pulmonary disease [COPD], diabetes, hypertension, and neoplasia). Clinical and laboratory data within the first 6 hours of ICU admission were also collected, such as mean arterial pressure (MAP), heart rate, respiratory rate, Glasgow Coma Scale (GCS) score, and arterial lactate levels.

Adverse events preceding AKI onset (e.g., hypotension, acute respiratory failure) were analyzed as risk factors, while events following AKI development (e.g., electrolyte imbalances such as hyperkalemia or hyponatremia) were included to evaluate complications. Notably, these events were the only variables with specific dates of occurrence beyond the time of patient admission, allowing temporal alignment with AKI development.

The primary outcome for this study was the development of AKI during the ICU stay, as documented in the patient’s medical records. AKI was considered present when explicitly listed in the problem list by the treating clinician. Although institutional guidelines incorporated assessments of serum creatinine and urine output per Kidney Disease: Improving Global Outcomes (KDIGO) criteria, the necessary variables for standardized AKI diagnosis were not available in the Epimed system [19]. Consequently, AKI diagnosis relied on clinician documentation.

### Statistical analysis

Descriptive statistics were employed to summarize patient characteristics, with categorical variables expressed as absolute and relative frequencies, and continuous variables as medians with interquartile range (IQR). Group differences in proportions and ranks were assessed using the chi-square test and Mann‒Whitney *U* test, respectively.

To assess potential risk factors, variables with clinical plausibility were included in a multivariable Cox regression model, using backwards stepwise method. Then, significant factors were incorporated into a Bayesian network model to visualize conditional dependencies between AKI and clinical or laboratory variables. Continuous variables were discretized using Hartemink’s algorithm via the “bnlearn” package, and the network structure was established with the Hill-Climbing algorithm to optimize conditional probability estimates [20–22]. Dependencies were represented as directed acyclic graphs, where each node corresponds to a variable and a direct arc between nodes represents a potential influence between variables. The robustness of the arcs was evaluated through non-parametric bootstrap testing with 100 replicates [23]. Arcs with more than 60% support were displayed.

For evaluating AKI-related complications, univariable Cox regression models were used for each event occurring after AKI development. Kaplan-Meier curves illustrated survival and hospital discharge probabilities, with log-rank tests used for group comparisons. Survival analysis over 6 weeks included an additional Cox regression model. Statistical significance was set at a p-value < 0.05. Analyses were performed using R, version 4.4.1 (The R Project for Statistical Computing, http://www.r-project.org/).

## RESULTS

### Study Population

During the study period, 6,563 patients were initially enrolled. Of these, 1,147 patients were excluded due to the presence of pre-existing CKD or with AKI at admission, or due to the missing of baseline or outcome data, leaving 5,416 patients in the final analysis (Fig. 1). A detailed comparison between excluded and included patients was provided in the supplementary material (S1 Appendix). Among the included patients, 369 (6.81%) patients developed AKI during their ICU stay, with a median time to development of 3 days (IQR: 1–5). The study cohort presented a median age of 69 years (IQR: 55–81), with a slight female predominance (3,032; 56%). Patients had a median SAPS3 score of 46 (IQR: 38–53), and most admissions were for non-surgical reasons (4,452; 82%) (Fig. 2A and Table 1).

**Fig. 1.**
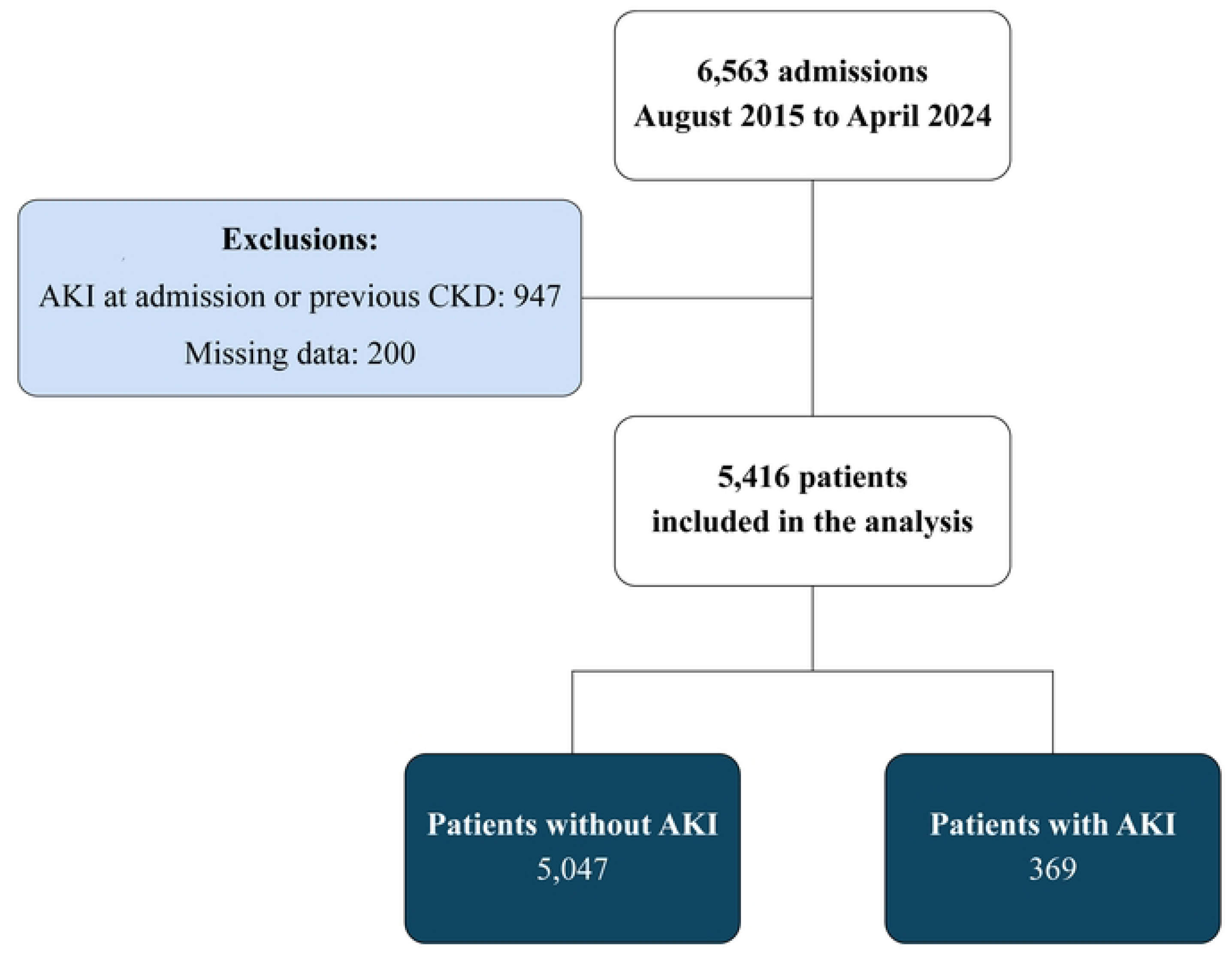
Study flowchart.

**Fig. 2.**
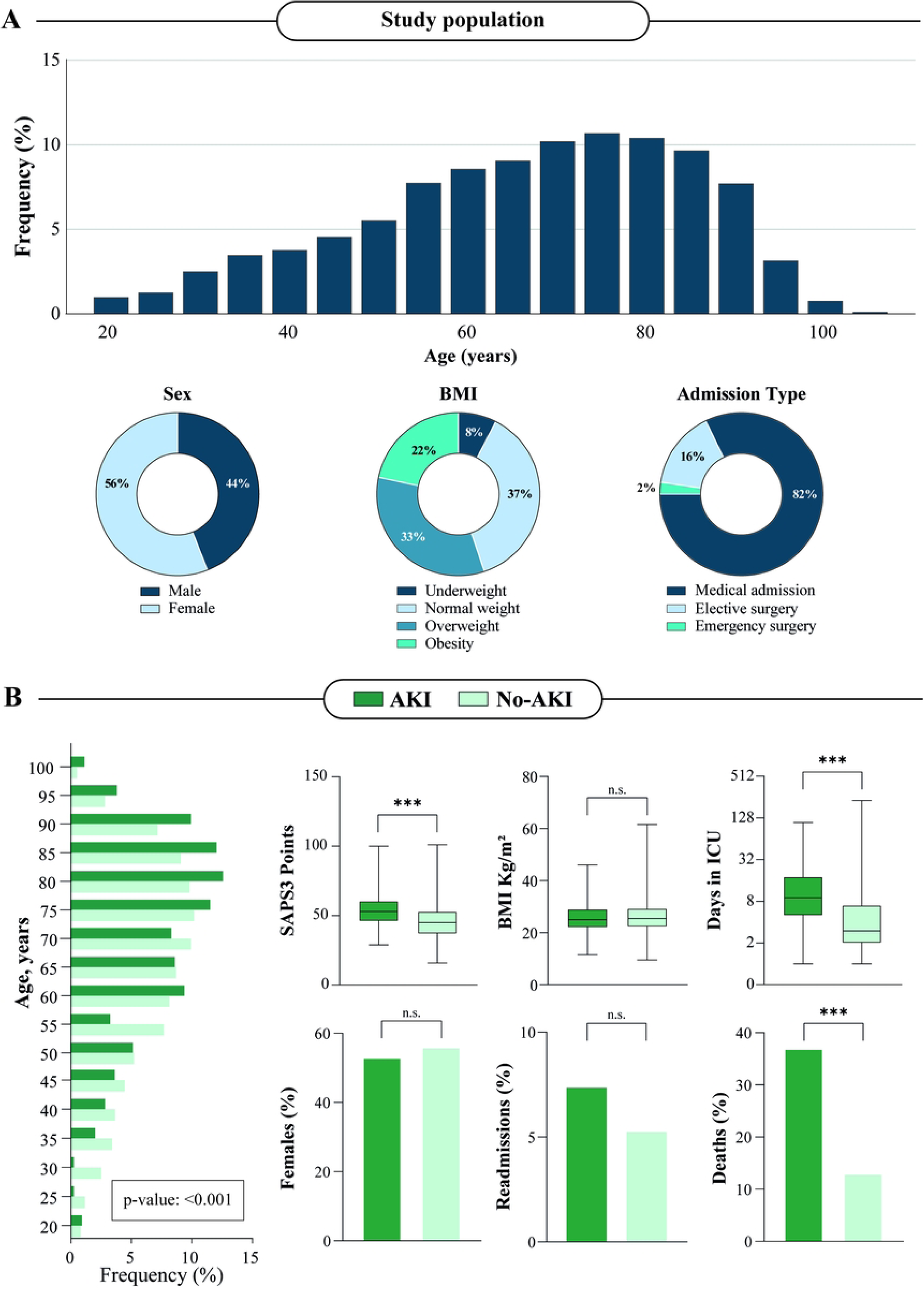
Population characteristics. (A) The upper panel presents a histogram illustrating the age distribution of the study participants at the baseline, while in the lower panel exhibits the distribution of sex, body mass index (BMI), and admission types among the study participants. (B) comparison between the groups of patients with (green) and without AKI (light green). To illustrate the values of p, the following symbols are used: p > 0.05 (ns), p ≤ 0.05 (*), p ≤ 0.01 (**), p ≤ 0.001 (***). A statistical significance level of p-value < 0.05 was adopted.

**Table 1.** Characteristics of ICU patients with and without acute kidney injury. Summarization of the clinical, demographic, and admission characteristics of patients included in the study, comparing those who developed AKI during their ICU stay to those who did not. Categorical variables are presented as absolute numbers (n) and percentages (%), while continuous variables are expressed as medians with interquartile ranges (IQR). Comparisons between groups were performed using the chi-square test for categorical variables and the Mann-Whitney U test for continuous variables. A p-value < 0.05 was considered statistically significant. Abbreviations: Acute Decompensated Heart Failure (ADHF); Acute Kidney Injury (AKI); Atrial Fibrillation (AFib); Body Mass Index (BMI); Chronic Obstructive Pulmonary Disease (COPD); Deep Vein Thrombosis (DVT); Glasgow Coma Scale (GCS); Intensive Care Unit (ICU); Interquartile Range (IQR); Mean Arterial Pressure (MAP); Myocardial Infarction (MI); Peripheral Vascular Disease (PVD); Simplified Acute Physiology Score 3 (SAPS3).

| Patient characteristics | Total (n = 5416) | No AKI (n = 5047) | AKI (n = 369) | p- value |
| --- | --- | --- | --- | --- |
| Age, years, median (IQR) | 69 (55 – 81) | 68 (54 – 80) | 74 (61 – 84) | <0.001 |
| Female, n (%) | 3032 (55.98) | 2834 (56.15) | 198 (53.66) | 0.380 |
| BMI, kg/m<sup>2</sup>, median (IQR) | 25.4 (22.3 – 29.3) | 25.5 (22.3 – 29.3) | 25 (22.1 – 29) | 0.633 |
| BMI categories |  |  |  |  |
| Underweight, n (%) | 410 (7.57) | 386 (7.65) | 24 (6.50) |  |
| Eutrophic, n (%) | 2018 (37.26) | 1861 (36.87) | 157 (42.55) |  |
| Overweight, n (%) | 1813 (33.47) | 1705 (33.78) | 108 (29.27) |  |
| Obesity, n (%) | 1175 (21.69) | 1095 (21.70) | 80 (21.68) |  |
| Admission type |  |  |  |  |
| Medical admission, n (%) | 4452 (82.20) | 4126 (81.75) | 326 (88.35) |  |
| Elective surgery, n (%) | 837 (15.45) | 803 (15.91) | 34 (9.21) |  |
| Emergency surgery, n (%) | 127 (2.34) | 118 (2.34) | 9 (2.44) |  |
| SAPS 3 score, median (IQR) | 46 (38 – 53) | 45 (37 – 53) | 53 (46 – 60) | <0.001 |
| Readmission, n (%) | 290 (5.35) | 263 (5.21) | 27 (7.32) | 0.106 |
| Deaths, n (%) | 797 (14.72) | 660 (13.08) | 137 (37.13) | <0.001 |
| Length of ICU stay, days, median (IQR) | 4 (2 – 7) | 3 (2 – 7) | 9 (5 – 18) | <0.001 |
| Length of hospital stay, days, median (IQR) | 8 (4 – 16) | 8 (4 – 15) | 16 (8 – 30) | <0.001 |
| Comorbidities |  |  |  |  |
| Heart failure, n (%) | 412 (7.61) | 383 (7.59) | 29 (7.86) | 0.930 |
| Chronic liver failure, *n* (%) | 20 (0.37) | 12 (0.24) | 8 (2.17) | <0.001 |
| Neoplasia, *n* (%) | 587 (10.84) | 543 (10.76) | 44 (11.92) | 0.543 |
| Immunosuppression, *n* (%) | 94 (1.74) | 83 (1.64) | 11 (2.98) | 0.091 |
| COPD, *n* (%) | 185 (3.42) | 167 (3.31) | 18 (4.88) | 0.146 |
| Hypertension, *n* (%) | 3673 (67.82) | 3395 (67.27) | 278 (75.34) | 0.002 |
| Asthma, *n* (%) | 176 (3.25) | 166 (3.29) | 10 (2.71) | 0.650 |
| Diabetes, *n* (%) | 1900 (35.08) | 1759 (34.85) | 141 (38.21) | 0.212 |
| Previous MI, *n* (%) | 416 (7.68) | 389 (7.71) | 27 (7.32) | 0.864 |
| Previous DVT, *n* (%) | 75 (1.38) | 72 (1.43) | 3 (0.81) | 0.457 |
| PVD, *n* (%) | 75 (1.38) | 67 (1.33) | 8 (2.17) | 0.270 |
| Chronic AFib, *n* (%) | 242 (4.47) | 219 (4.34) | 23 (6.23) | 0.117 |
| Previous stroke, *n* (%) | 778 (14.36) | 724 (14.34) | 54 (14.63) | 0.939 |
| Smoking, *n* (%) | 369 (6.81) | 344 (6.82) | 25 (6.77) | 1.000 |
| Alcoholism, *n* (%) | 219 (4.04) | 197 (3.90) | 22 (5.96) | 0.072 |
| Peptic ulcer disease, *n* (%) | 24 (0.44) | 21 (0.42) | 3 (0.81) | 0.483 |
| Thyroid disorder, *n* (%) | 377 (6.96) | 345 (6.84) | 32 (8.67) | 0.218 |
| Dyslipidemia, *n* (%) | 492 (9.08) | 457 (9.05) | 35 (9.48) | 0.854 |
| Adverse events |  |  |  |  |
| Hypothermia, *n* (%) | 407 (7.5) | 369 (7.3) | 38 (10.3) | 0.041 |
| Hypotension, *n* (%) | 1044 (19.3) | 929 (18.4) | 115 (31.2) | <0.001 |
| Acute respiratory failure, *n* (%) | 899 (16.6) | 810 (16.0) | 89 (24.1) | <0.001 |
| ADHF, *n* (%) | 304 (5.6) | 277 (5.5) | 27 (7.3) | 0.091 |
| Hyperkalemia, *n* (%) | 155 (2.86) | 97 (1.92) | 58 (15.72) | <0.001 |
| Hypokalemia, *n* (%) | 628 (11.59) | 540 (10.70) | 88 (23.85) | <0.001 |
| Hypernatremia, *n* (%) | 219 (4.04) | 160 (3.17) | 59 (15.99) | <0.001 |
| Hyponatremia, *n* (%) | 212 (3.91) | 182 (3.61) | 30 (8.13) | <0.001 |
| Hypocalcemia, *n* (%) | 155 (2.86) | 131 (2.60) | 24 (6.50) | <0.001 |
| Admission parameters |  |  |  |  |
| Lowest MAP, mmHg, median (IQR) | 96.7 (83.3 – 110.6) | 96.7 (83.3 – 110.7) | 96.7 (81.0 – 111.0) | 0.525 |
| Highest Heart Rate, bpm, median (IQR) | 84 (71 – 98) | 83 (71 – 97) | 89 (75 – 104.50) | <0.001 |
| Highest Respiratory Rate, ipm, median (IQR) | 20 (18 – 22) | 20 (18 – 22) | 20 (18 – 24) | <0.001 |
| Lowest GCS, points, median (IQR) | 15 (13 – 15) | 15 (13.5 – 15) | 14 (12 – 15) | <0.001 |
| Highest Arterial Lactate, mmol, median (IQR) | 1.5 (1.1 – 2.24) | 1.5 (1.1 – 2.2) | 1.7 (1.2 – 2.7) | <0.001 |

### Baseline Characteristics and Risk Factors for AKI

Patients with AKI were significantly older, with a median age of 74 years (IQR: 61-84) compared to 68 years (IQR: 54–80; p-value <0.001) in those without AKI. They were more likely to have comorbidities, such as hypertension (75.34% vs. 67.27%, p-value = 0.002) and chronic liver failure (2.17% vs. 0.24%, p-value < 0.001) (Figure 2B). No differences were observed in sex proportions, BMI categories, or smoking status between the two groups. AKI patients had longer hospital stay (median 16 vs. 10 days, p-value < 0.001) and higher mortality rates (37.13% vs. 10.54%, p-value < 0.001).

At ICU admission, AKI patients exhibited higher arterial lactate (1.7 mmol/L, IQR: 1.2–2.7 vs. 1.5 mmol/L, IQR: 1.1–2.2, p-value < 0.001), respiratory rate (20 ipm, IQR: 18–24 vs. 20 ipm, IQR: 18–22, p-value < 0.001), heart rate (89 bpm, IQR: 75–104 vs. 83 bpm, IQR: 71–97, p-value < 0.001) and lower Glasgow coma scale points (14, IQR: 12–15, vs. 15, IQR: 13.5–15, p-value < 0.001). Additionally, adverse events, occurring prior to AKI development, including hypotension, hypothermia, and acute respiratory failure, were significantly more common in AKI patients.

Multivariable Cox regression analysis identified age (adjusted Hazard Ratio [aHR] 1.01; 95% CI: 1.00–1.02; p-value = 0.008), chronic liver failure (aHR 10.02; 95% CI: 4.91–20.41; p-value < 0.001), hypertension (aHR 1.29; 95% CI: 1.00–1.66; p-value = 0.048), heart rate at admission (aHR 1.01; 95% CI: 1.00–1.01; p-value = 0.004), and reduced level of consciousness (aHR 1.67 per point decrease; 95% CI: 1.34–2.07; p-value < 0.001) as independent predictors of AKI. Underweight patients had a reduced risk of AKI compared to those with normal weight (aHR 0.59; 95% CI: 0.38–0.91; p-value = 0.017), while no significant associations were observed for overweight or obese patients (Figure 3).

**Figure 3.**
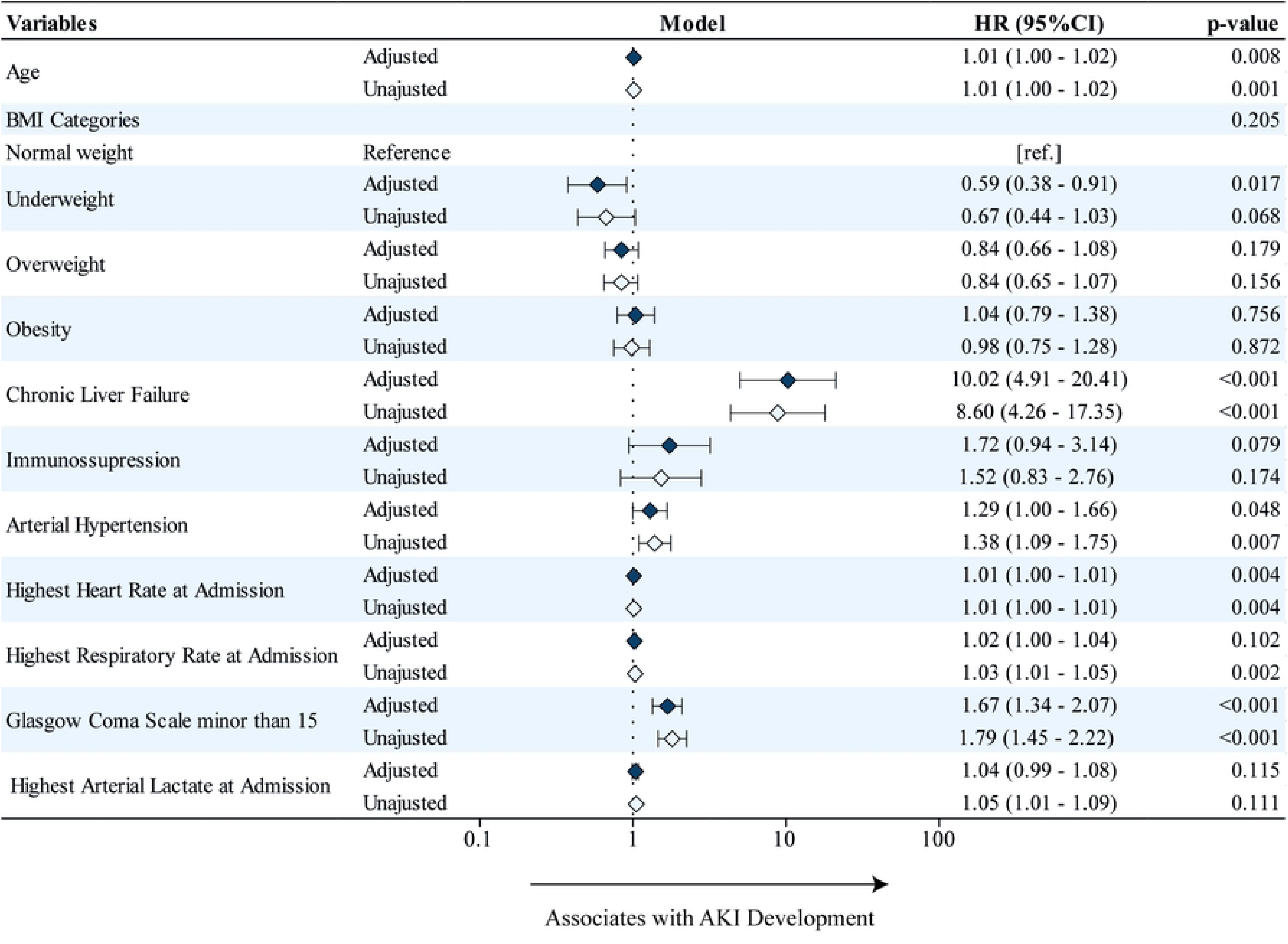
Adjusted and unadjusted Cox regression models for AKI development. Forest plot presenting the adjusted (dark blue) and unadjusted (light blue) hazard ratios with 95% confidence intervals for variables associated with the development of acute kidney injury (AKI) in intensive care unit (ICU) patients. The dotted line represents the null point (HR = 1). A p-value < 0.05 represents statistical significance.

The Bayesian network model highlighted strong conditional dependencies between AKI and reduced level of consciousness at admission and chronic liver failure (Figure 4). Additionally, age was indirectly linked to AKI through strong associations with arterial hypertension, BMI, and elevated heart rate at admission.

**Figure. 4.**
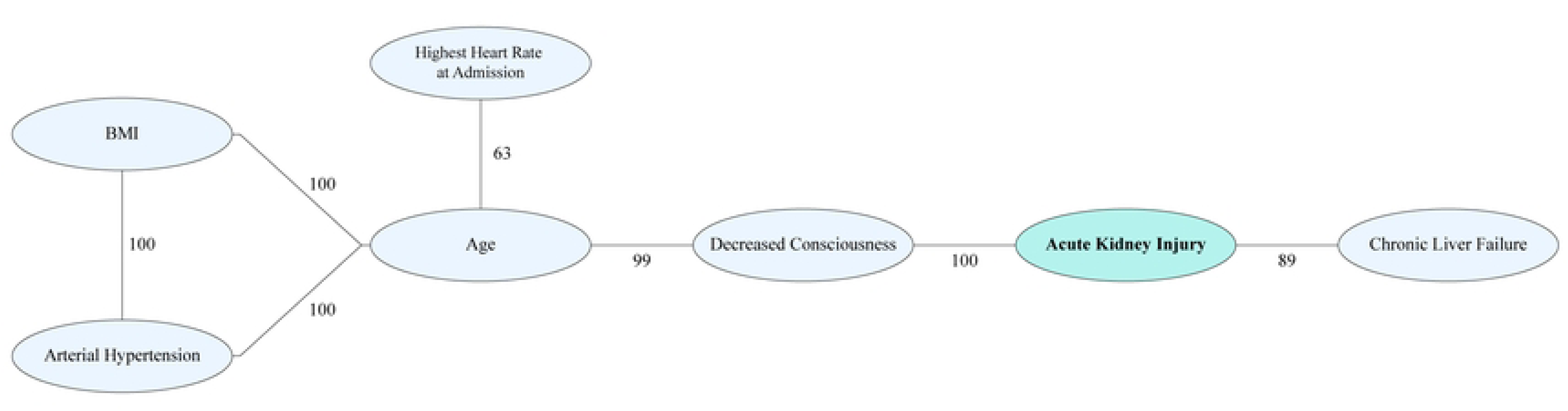
Bayesian network to describe conditional dependencies between the AKI and statistically significant risk factors. The Bayesian network model with bootstrap (100x) illustrates the dependencies between acute kidney injury (AKI) and statistically significant risk factors. Direct associations are represented by lines. Associations that remained statistically significant on >60 times out of 100 bootstraps are plotted, and the numbers of times each association persisted during bootstrap are shown.

### Complications Associated with AKI Development

Electrolyte disturbances were significantly more frequent in patients who developed AKI during their ICU stay, including hyperkalemia (HR 2.94; 95% CI: 2.00–4.33; p-value < 0.001), hypokalemia (HR 1.76; 95% CI: 1.23–2.53; p-value < 0.001), hypernatremia (HR 1.73; 95% CI: 1.23–2.44; p-value: < 0.001), hyponatremia (HR 1.53; 95% CI: 1.14–2.06; p-value < 0.001), and hypocalcemia (HR 1.49; 95% CI: 1.07– 2.06; p-value = 0.018).

The Kaplan-Meier curves of survival over a 6-week period demonstrated that patients who developed AKI exhibited shorter survival over time (HR 1.56; 95% CI: 1.29–1.88; p-value: < 0.001) (Figure 5A). Furthermore, the Kaplan-Meier curves for hospital length of stay revealed that AKI patients had a significantly higher probability of prolonged hospitalization (p-value: < 0.001) (Figure 5B).

**Figure. 5.**
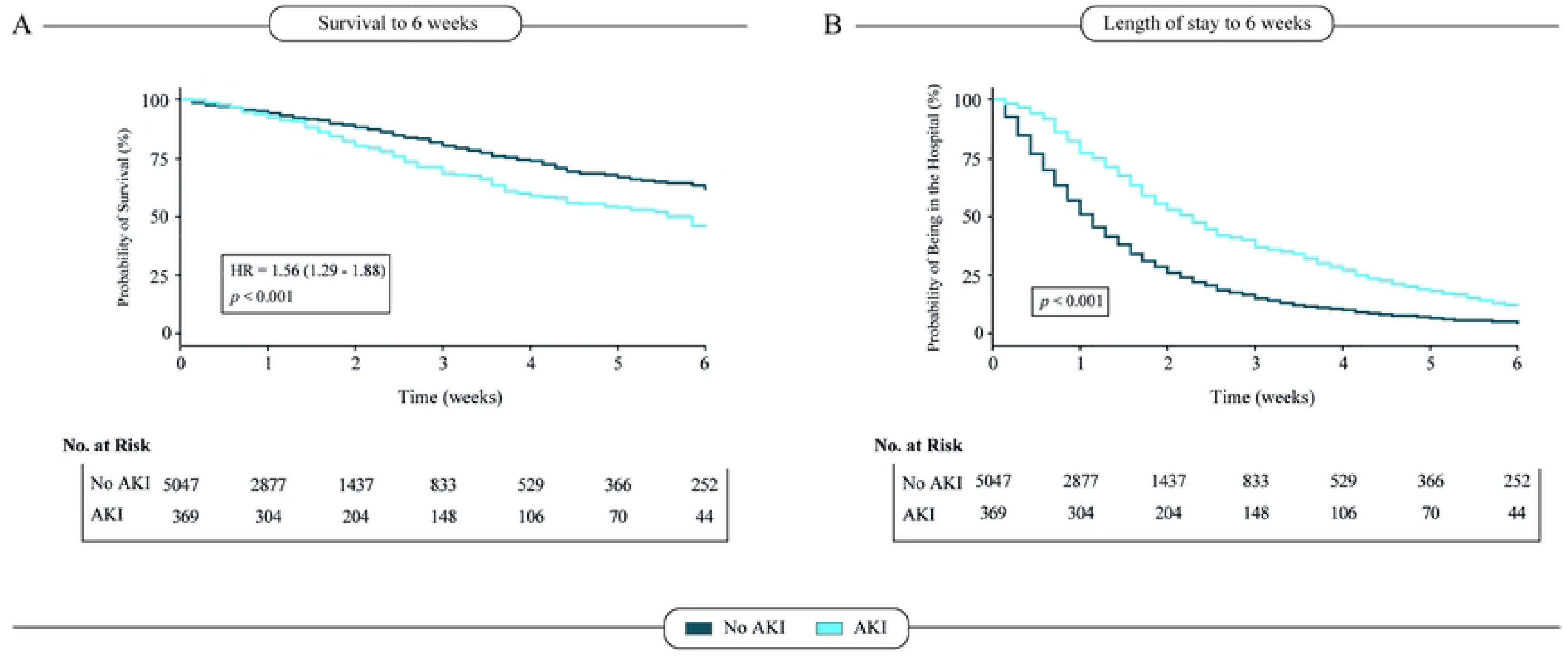
Kaplan-Meier Analysis of AKI Survival and Hospital Length of Stay. (A) Kaplan-Meier curves of survival to 6 weeks among the patients in both groups, with significantly higher mortality among those with acute kidney injury (AKI) in the intensive care unit (ICU). (B) illustrates Kaplan-Meier curves of the probability of remaining in the hospital according to the clinical distinction of No AKI vs AKI.

## DISCUSSION

This retrospective cohort study identified significant clinical parameters associated with ICU-acquired AKI in a LMIC setting. Our findings highlight the importance of age, chronic liver failure, arterial hypertension, heart rate, and reduced levels of consciousness as independent predictors of AKI, with potential implications for early detection and intervention to mitigate its impact.

The incidence of AKI in this cohort (6.81%) is significantly lower than rates reported in both high-resource settings (25–30%) and other resource-limited settings, where incidences of approximately 14% have been observed [2,3,14,17]. This discrepancy may be attributed to differences in the socioeconomic status of patient populations, illness severity, hospital setting, and diagnostic practices. In our study, AKI diagnosis relied on clinician documentation without standardized criteria due to limitations in available data. This may have led to under-recognition of AKI cases. Additionally, the patient population in this Brazilian ICU presented lower general severity compared to cohorts in other studies, potentially contributing to the lower AKI incidence [24].

Age was significantly associated with an increased hazard of AKI, consistent with global literature linking advanced age to declining renal resilience and comorbidities such as hypertension and chronic liver failure [9–11]. Renal involvement tends to increase throughout life due to cumulative exposure to nephrotoxic medications, lifestyle habits, and other stressors [25,26]. Notably, the Bayesian network underscored the indirect role of age in AKI development through its association with these comorbidities and hemodynamic parameters, highlighting the multifactorial nature of AKI risk.

Hypertension was one of the most prevalent comorbidities in this cohort and was significantly associated with AKI development. This finding aligns with prior studies linking elevated blood pressure to kidney injury through mechanisms such as oxidative stress, chronic inflammation, and mechanical strain on renal vasculature [25,27].

Furthermore, chronic liver failure emerged as the strongest independent predictor of AKI, with a tenfold increased hazard and a direct relationship, as demonstrated by the Bayesian network analysis. This result aligns with previous studies highlighting the profound impact of liver dysfunction on renal outcomes [28]. While hepatorenal syndrome (HRS) was not directly evaluated, its pathophysiological features, including altered renal perfusion and systemic inflammation, likely contributed to the observed association [29]. These associations suggest that the severity of comorbidities may contribute to the vulnerability of patients to AKI [30].

Regarding clinical parameters, patients who developed AKI exhibited higher heart rates and reduced levels of consciousness at admission, both of which were independently associated with AKI. Although limited studies have directly examined these variables in relation to AKI, they are widely recognized as critical markers of illness severity [31,32]. Elevated heart rate may reflect underlying hemodynamic instability, such as hypovolemia or sepsis, which compromises renal perfusion. Similarly, reduced Glasgow Coma Scale scores, a key component of prognostic scores like SOFA and SAPS 3, may indicate systemic dysfunction or the need for sedatives and mechanical ventilation, both of which are potential contributors to renal injury [33,34].

Interestingly, underweight patients had a lower risk of developing AKI compared to those with normal weight, while no significant differences were found for overweight or obese patients. This finding contrasts with studies suggesting obesity as a risk factor for AKI [35,36]. However, previous study highlighted that, particularly in older patients, underweight was associated with AKI development through possible diminished physiological reserves [37]. These parallels highlight the complex interplay between nutritional status, metabolic reserves, and AKI risk, underscoring the importance of considering population-specific characteristics and malnutrition as a potentially underrecognized contributor to AKI.

AKI was associated with significant complications, including electrolyte imbalances such as hyperkalemia and hypernatremia, which are known to exacerbate morbidity and mortality in ICU patients. Kidney injury has been demonstrated to result in an important electrolyte and metabolic imbalances, which can lead to complications such as arrhythmias [38–40]. Furthermore, patients who developed AKI exhibited poorer outcomes, including reduced survival and prolonged hospitalization. These findings are already well established in the literature and underscore the impact of this condition on ICU prognosis [41,42].

This study has several limitations The retrospective design and reliance on clinician-documented AKI without standardized diagnostic criteria may have led to underestimation of AKI incidence and misclassification bias. The necessary variables for applying standardized AKI criteria (e.g., serial serum creatinine measurements, urine output) were not available in the Epimed system. Additionally, data on important risk factors such as nephrotoxic medication exposure, sepsis, contrast agent use, and fluid balance were not available, limiting the ability to adjust for these confounders. Additionally, data entry was performed by ICU nurses, which may introduce data quality concerns despite standardized protocols. The study was conducted in a single center, limiting generalizability to other settings. Despite these limitations, the results provide valuable insights into AKI risk factors and outcomes in a resource-limited setting.

This study identified advanced age, chronic liver failure, arterial hypertension, elevated heart rate at admission, and reduced level of consciousness as independent risk factors for the development of AKI in critically ill patients admitted to a Brazilian ICU. AKI was associated with higher mortality and prolonged hospitalization, emphasizing the importance of early identification and targeted interventions. Recognizing these risk factors can aid clinicians in implementing preventive strategies and optimizing patient management, ultimately improving outcomes in this vulnerable population.

## Data Availability

Data will be available upon reasonable request to the corresponding author.

## DECLARATIONS

### Ethical approval and consent to participate

All the clinical aspects were evaluated according to the principles of Resolution 466/2012 of the National Health Council and the Helsinki Declaration. The research project was approved by the Ethics and Research Committee of the Salvador University under CAAE number (61562222.2.0000.5033), with patient consent waived.

## Consent for publication

Not applicable.

## Availability of data and materials

The datasets used and analyzed during the current study are available from the corresponding authors on reasonable request.

## Authors’ contributions

LC-M, RM-C, RM-T and IB-F: conceptualization, design of study, and manuscript draft. RM-C and RM-T: investigation and visualization. RM-C: data acquisition. RM-T: data analysis and interpretation. RM-C and BA: supervision, critical revision, editing and final approval of the manuscript. All the authors have given final approval for the version of the manuscript to be published.

## Acknowledgements

The authors thank those who contributed directly or indirectly to the construction of this article, research groups GEMINI, linked to the Núcleo de Ensino e Pesquisa do Hospital da Cidade, and the MONSTER Institute.

